# The role of the environment in overweight and eating behavior variability: Insights from a multivariate twin study

**DOI:** 10.1101/2020.03.23.20041384

**Authors:** Moritz Herle, Juan J Madrid-Valero, José J Morosoli, Lucía Colodro-Conde, Juan Ordoñana

## Abstract

The health consequences of overweight and obesity remain one of the greatest global health challenges. Twin and molecular studies have confirmed the genetic basis of individual differences in BMI; however, genetics cannot explain the rapid rise of obesity rates over the past decades. Eating behaviors have been stipulated to be the behavioral expression of genetic risk in an obesogenic environment. Multivariate twin studies can inform future applied research by providing insights into the etiology of the relationship between behaviors and BMI. In this study, we aimed to decompose variation and covariation between three key eating behaviors and BMI in a sample from a population-based twin registry of adult women in the southeast of Spain, The Murcia Twin Registry (345 complete, 9 incomplete, same-sex female twin pairs, 175 MZ, 170 DZ). Phenotypes were emotional eating, uncontrolled eating, and cognitive restraint, as measured by the Three Factor Eating Questionnaire, as well as objectively measured BMI. Variation in eating behaviors was mostly driven by non-shared environmental factors (range: 56-65%), whereas shared environmental and genetic factors were secondary. All three eating behaviors were associated with BMI at a phenotypic level (range r=0.19–0.25). Etiological correlations implied that non-shared environmental factors underly the covariations of the phenotypes (Emotional eating – Uncontrolled eating: rE= 0.54, 95%CI: 0.43, 0.64; BMI – Cognitive restraint: rE= 0.15, 95%CI: 0.01, 0.28). Results indicate that in contrast to BMI, individual differences in eating behaviors are mostly explained by non-shared environmental factors, which also account for the phenotypic correlation between eating behaviors and BMI. These results support that eating behaviors are viable intervention targets to help individuals reach and maintain a healthy weight.

## Introduction

Obesity rates remain high, with 28% of adults classifying as having overweight or obesity worldwide (M. Ng et al., 2014). This is alarming as overweight and obesity have been associated with negative health consequences including cardiovascular disease, cancer and overall mortality (Abdelaal, le Roux, & Docherty, 2017; Dixon, 2010). However, management of overweight and obesity requires understanding of the intricate interactions of genetic, physiological, behavioral, and social factors that drive body weight (Bray et al., 2018).

In recent years, the genetic basis of obesity has been supported by large-scale genome-wide studies, detecting 97 genetic variants associated with body mass index (BMI) (Locke et al., 2015). Despite the genetic contribution to obesity, genetic factors cannot account for its rapid rise of over the past decades. Environmental factors such as greater portion sizes (Piernas & Popkin, 2011; Zlatevska, Dubelaar, & Holden, 2014) of high fat foods (Swinburn et al., 2011), increased sedentary work and leisure time (S. W. Ng & Popkin, 2012), and abandoning traditional dietary habits (Leon-Munoz et al., 2012) have all been proposed to contribute to the obesogenic environment resulting in high rates of obesity. However, regardless of the pervasiveness of these environmental factors, not everyone has overweight or obesity, and we observe large individual differences in weight in the population.

The behavioral susceptibility theory of obesity (C. Llewellyn & Wardle, 2015) aims to explain this dual influence of genes and environments by hypothesizing that eating behaviors are the behavioral expression of a genetic susceptibility for obesity in response to an obesogenic food environment. Previous studies have highlighted that eating behaviors, such as food-cue responsiveness and emotional overeating, the tendency to eat in response to negative emotion, such as sadness or loneliness, are associated with body weight and food intake in cross-sectional (Angle et al., 2009; Hunot, Fildes, Croker, Llewellyn, et al., 2016; van den Boer et al., 2017) and longitudinal studies (Syrad, Johnson, Wardle, & Llewellyn, 2016; van Jaarsveld, Boniface, Llewellyn, & Wardle, 2014). However, less is known about the etiology of eating behaviors.

Twin studies, conducted to tease apart the genetic and environmental contributions to individual differences in eating behaviors, have been used to estimate the heritability and environmental influences of eating behaviors in childhood (C. H. Llewellyn, C. H. M. van Jaarsveld, L. Johnson, S. Carnell, & J. Wardle, 2010b). Reported estimates of heritability (the relative contribution of genetic factors to individual differences) range widely: from 5% for emotional over- and undereating in childhood (M. Herle, Fildes, & Llewellyn, 2018; Moritz Herle, Fildes, Steinsbekk, Rijsdijk, & Llewellyn, 2017), to 72% for satiety responsiveness in infancy (C. H. Llewellyn, C. H. van Jaarsveld, L. Johnson, S. Carnell, & J. Wardle, 2010a). In addition, research in children has proposed that increased genetic risk for obesity was associated with decreased responsiveness to internal satiety cues, supporting the behavioral susceptibility theory of obesity (C. H. Llewellyn, Trzaskowski, van Jaarsveld, Plomin, & Wardle, 2014). In adults, four studies have estimated the heritability of eating behaviors in Swedish (Tholin, Rasmussen, Tynelius, & Karlsson, 2005), UK and Finnish (Keskitalo et al., 2008), Korean (Sung, Lee, Song, Lee, & Lee, 2010), and Sri Lankan twins (M. P. Herle et al., 2019). Produced estimates vary widely across studies: 9-72% for emotional eating, 45-77% for responsiveness to external food cues, and 31-69% for restraint or fasting behaviors (M. P. Herle et al., 2019; Keskitalo et al., 2008; Sung et al., 2010; Tholin et al., 2005). Only one study examined the shared etiology of eating behaviors and BMI, with some evidence for shared genetic influences (Keskitalo et al., 2008).

These heterogeneous results might be explained by differences between the studied populations. Previous twin studies included individuals with a large age range (late adolescence to older adulthood), were often drawn from public health registers, coming from markedly different regions and socio-economic background within each country. This is important, because, in the case of BMI, genetic factors have been found to contribute more to the differences in BMI in twins of highly educated parents, whereas environmental factors were found to be more important in families of parents with limited education (Lajunen, Kaprio, Rose, Pulkkinen, & Silventoinen, 2012), which is congruent with recent evidence for gene-environment interactions for BMI (Komulainen et al., 2018).

In the current study, we aim to address some of the drawbacks of previous research by using a distinctive sample of women in the middle adulthood, with a shorter age range, from a smaller geographical area, and with a regional specific food environment (southeast of Spain). We aim to shed light on specific areas that have not been explored sufficiently in the literature, such as the nature of relationship between BMI and eating behaviors, the contribution of genetic and environmental factors in a female adult sample, and the expression of genetic susceptibility to obesity in a Mediterranean country.

## Methods

### Participants

Data came from the Murcia Twin Registry (MTR), a population-based twin registry of adult multiples born between 1940 and 1976 in the region of Murcia, Southeast of Spain. Information regarding the MTR characteristics and recruitment procedures can be found elsewhere (Ordonana et al., 2019). Being a population-based registry with active recruitment, MTR participants are fairly representative of the general population, of the corresponding ages, in terms of demographic and health data. A comparison with reference surveys at a regional and national level showed that MTR participants presented comparable prevalence in chronic conditions and other health variables as the general population of Murcia and Spain (Ordonana et al., 2018). The sample for this study was composed of 345 complete (9 incomplete) same-sex female twin pairs (175 MZ; 170 DZ), born between 1946 and 1966, who participated in the data collection in 2009 (age at data collection: mean = 53.13, SD = 7.55, range 43 - 69). All registry and data collection procedures involved in this study were approved by the Murcia University Ethics Committee, and informed consent was obtained from all twins.

### Measures

Eating behaviors were measured with the Spanish version of the Three Factor Eating Behavior Questionnaire – R18 (TFEQ-R) (Jauregui-Lobera, Garcia-Cruz, Carbonero-Carreno, Magallares, & Ruiz-Prieto, 2014) which is a shortened version of the original TFEQ (Stunkard & Messick, 1985). The questionnaire consists of three subscales: Emotional eating (three items, example: “When I feel lonely, I console myself with eating”), uncontrolled eating (nine items, example: “Sometimes when I start eating, I just can’t seem to stop”), and cognitive restraint (six items, example: “I consciously hold back at meals in order not to gain weight”). The three subscales had good internal consistency in this sample: Emotional eating, McDonald’s omega= 0.77 (95%CI:0.61, 0.86); Uncontrolled eating, McDonald’s omega =0.9 (95% CI: 0.82, 0.94) and Cognitive restraint, McDonald’s omega =0.86 (95% CI:0.80, 0.92).

A blinded research assistant collected anthropometric measurements during data collection interviews. Body weight was measured using a TANITA BC-240 MA (Tanita Corporaton of America, USA) and height using a portable stadiometer. BMI was calculated by dividing the individuals’ body weight in kilograms by the square of their height in meters.

### Analyses

The twin design is based on the comparison of monozygotic (MZ) and dizygotic (DZ) twins. MZ twins share 100% of their DNA, whereas DZ twins share on average about half of their segregating genes. Importantly, as both types of twins are exposed to very similar environments, such as intrauterine exposures and aspects of parental upbringing, difference in the correlation between MZ and DZ twin pairs are assumed to reflect genetic differences (Boomsma, Busjahn, & Peltonen, 2002). These genetic components can be divided into additive (A; i.e., summed allelic effects across multiple genes) and non-additive (D; i.e., genetic dominance, possibly including epistasis) factors, whereas the environmental factors can be divided into shared (C; i.e., common/family environment) and individual or unique (E; i.e., idiosyncratic experiences, including measurement error) environmental factors. It is not possible to estimate C and D simultaneously in a classical twin model, and the choice of modelling C or D depends on the pattern of MZ and DZ correlations. Usually C is estimated if the DZ twin correlation is more than half of the MZ twin correlation, and D is estimated if the DZ twin correlation is less than half of the MZ correlation (Grasby, Verweij, Mosing, Zietsch, & Medland, 2017; Rijsdijk & Sham, 2002). Overall, a greater phenotypic similarity in MZ twin pairs compared with DZ twin pairs is attributed to genetic influences (A or D components), under the assumption that both MZ and DZ twins are exposed to equal shared environments (38). As an extension to this concept, multivariate twin models decompose in addition the covariance between multiple phenotypes following the same principles. A multivariate ACE model provides etiological correlations (denoted rA, rC, and rE, ranging from −1 to 1) which indicate the extent to which the A, C, and E factors that explain individual differences in one phenotype also affect the other. Furthermore, the derived etiological correlations can be used to decompose the phenotypic correlation between the two phenotypes: by dividing the etiological correlations by the phenotypic correlation, the proportion of the correlation that is due to A, C and E can be calculated (Rijsdijk & Sham, 2002). Maximum likelihood structural equation modelling is used to fit the models and provide parameter estimates with 95% confidence intervals. Model fit is compared using likelihood ratio test (Rijsdijk & Sham, 2002) and the Akaike Information Criterion (AIC) (Posada & Buckley, 2004). First, a saturated model with no parameter constraints is fitted. Assumptions of the twin design are also tested, including the homogeneity of the means and variances of first and second born twins, and across zygosity groups. Followed by the ACE model, constraining the A and C paths to their theoretical values within MZ and DZ pairs (1 and 0.5). Further details of the twin design, including testing assumptions, can be found elsewhere (Posthuma et al., 2003). In this study, a multivariate ACE model including the eating behavior variables and BMI was fit.

Prior to structural equation analyses, mean scores were calculated for EE, UE and CR. Only participants who answered all items of the questionnaire were included. Scores were regressed on age at measurement as this is shared completely within twin pairs and potentially inflate twin-pair similarity (Mcgue & Bouchard, 1984). All analyses were conducted in R using the OpenMx package (Boker et al., 2011). The analytical code is available on https://github.com/MoritzHerle/The-role-of-the-environment-in-overweight-and-eating-behavior-variability.

## Results

Descriptive statistics can be found in Table 1. Two thirds of the participants had either overweight or obesity.

**Table 1.** Descriptive statistics for analyses sample (total N= 699)

| **Zygosity** | **N (%) or Mean (SD)** |
| --- | --- |
| Monozygotic | 175 pairs |
| Dizygotic | 170 pairs |
| **Age** | 53.13 (7.55) |
| **Education** |  |
| Illiterate | 2 (<1) |
| No education (reading & writing) | 72 (10) |
| Primary school | 239 (34) |
| Secondary school | 187 (27) |
| Vocational training | 65 (9) |
| A-Levels | 38 (5) |
| University degree | 60 (13) |
| Missing | 17 (2) |
| **Body mass index** | 27.58 (5.06) |
| Healthy weight, 18.5 - 25 | 228 (32) |
| Underweight, <18 | 5 (1) |
| Overweight, 25-30 | 264 (38) |
| Obese, >30 | 197 (28) |
| Missing | 5 (1) |
| **Emotional eating** | 1.60 (0.80) |
| **Cognitive restraint** | 2.24 (0.94) |
| **Uncontrolled eating** | 1.49 (0.56) |

### Phenotypic correlations

All included phenotypes were positively correlated. Highest correlation was found between emotional eating and uncontrolled eating, see Table 2 for all correlations.

**Table 2.** Phenotypic correlations (95% Confidence intervals) of eating behaviors and body mass index in the Murcia twin register (N=699)

|  | **EE** | **CR** | **UE** | **BMI** |
| --- | --- | --- | --- | --- |
| **EE** | 1 |  |  |  |
| **CR** | 0.16 (0.08, 0.24) | 1 |  |  |
| **UE** | 0.59 (0.53, 0.64) | 0.12 (0.04, 0.20) | 1 |  |
| **BMI** | 0.19 (0.11, 0.27) | 0.25 (0.17, 0.32) | 0.20 (0.12, 0.28) | 1 |

Abbreviations: EE: Emotional eating; CR: Cognitive restraint; UE: Uncontrolled eating; BMI: Body mass index

Intraclass correlations indicate the similarity of one twin with their co-twin and are compared across MZ and DZ twin pairs. With the exception of cognitive restraint, MZ twin intraclass correlations were greater (but not more than twice) than DZ intraclass correlations, indicating that a model including additive genetic and common environment effects might be the most suitable for the data, instead of assuming additive and dominant genetic effects (Rijsdijk & Sham, 2002). See Table 3 for all intraclass correlations.

**Table 3.** Intraclass correlations (95% Confidence intervals) of eating behaviors and body mass index for monozygotic (MZ) and di-zygotic (DZ) twin pairs (N=699)

|  | **MZ**<br>(N=175 pairs) | **DZ**<br>(N=170 pairs) |
| --- | --- | --- |
| **EE** | 0.35 (0.21, 0.48) | 0.22 (0.05, 0.37) |
| **CR** | 0.32 (0.17, 0.45) | 0.34 (0.18, 0.47) |
| **UE** | 0.38 (0.24, 0.50) | 0.36 (0.20, 0.50) |
| **BMI** | 0.66 (0.56, 0.73) | 0.39 (0.23, 0.52) |

Abbreviations: MZ: Monozygotic twins; DZ: Dizygotic twins; EE: Emotional eating; CR: Cognitive restraint; UE: Uncontrolled eating; BMI: Body mass index

### Decomposition of variance

Means and variances of all variables could be equated across twin order and zygosity groups, suggesting that assumptions of the twin design were met.

A multivariate ACE model (including all parameters - A, C and E - for BMI and all three eating behaviors, as well as rA, rC and rE between them) with means and variance equated within first and second born twins and across zygosity groups fit the data well as the comparison with the saturated model indicated no significant drop in model fit between the two models (χ^2^ = 63.130, *p*=0.18). In line with likelihood ratio test, AIC also favored the more parsimonious multivariate ACE model over the saturated model (AIC_ACE_: 12203.6, vs. AIC_FULL_: 12248.5). Full fit statistics for the saturated and multivariate ACE model are shown in Supplementary Table 1.

**Supplementary Table 1.** Fit statistics for twin model fitting, Murcia twin registry (N=699)

| Model | ep | df | -2LL | AIC | $\Delta \chi^2$ (df) | *p-value* |
| --- | --- | --- | --- | --- | --- | --- |
| Full Saturated | 88 | 2634 | 17516.51 | 12248.51 |  |  |
| ACE | 34 | 2692 | 17579.64 | 12203.6 | 63.130 (54) | 0.18 |

Abbreviations: ep: estimated parameters; df: degrees of freedom; -2LL: -2 log-likelihood; AIC: Akaike's Information Criterion, $\Delta \chi^2$ : differenced in chi-square

Table 4 shows the parameter estimates for A, C and E (and 95% CI), indicating the relative importance of genetic, shared environmental, and non-shared environmental influences (including measurement error) on variation in BMI and the three eating behaviors. Variation in all eating behaviors was mostly explained by non-shared environmental factors (range from 0.56 – 0.64). Genetics and shared environment were of less importance, except for cognitive restraint, for which shared environmental factors accounted for about one quarter of the variance (C=0.26, 95%CI: 0.04, 0.40). In contrast, variation in BMI was similarly explained by genetic effect (A= 0.43, 95%CI: 0.13, 0.69) and non-shared environmental effects (E=0.36, 95%CI: 0.29, 0.45).

**Table 4.** Results from multivariate ACE model, univariate variance components (N=699)

|  | **A** | **C** | **E** |
| --- | --- | --- | --- |
| **EE** | 0.26 (0.00, 0.47) | 0.10 (0.00, 0.35) | 0.64 (0.52, 0.78) |
| **CR** | 0.09 (0.00, 0.34) | 0.26 (0.04, 0.40) | 0.65 (0.55, 0.76) |
| **UE** | 0.22 (0.00, 0.53) | 0.22 (0.00, 0.45) | 0.56 (0.45, 0.70) |
| **BMI** | 0.43 (0.13, 0.69) | 0.21 (0.00, 0.47) | 0.36 (0.29, 0.45) |

Abbreviations: A: Additive genetic factors, C: shared environmental factors, E: non-shared environmental factors, EE: Emotional eating; CR: Cognitive restraint; UE: Uncontrolled eating; BMI: Body mass index

### Decomposition of Covariance

As shown in Table 2, all variables were significantly positively correlated with each other. Most of the etiological correlations (rA, rC, and rE) were non-significant, with large confidence intervals often including ‘0’, likely due to low statistical power (see Table 5). Uncontrolled eating and emotional eating were found to be influenced, in part, by similar non-shared environmental factors (rE=0.54, 95%CI: 0.43, 0.64). Furthermore, results indicated that cognitive restraint and BMI had non-shared environmental factors in common (rE=0.15, 95%CI: 0.01, 0.28), as well as uncontrolled eating and BMI (rE=0.15, 95%CI: 0.00, 0.29).

**Table 5.**
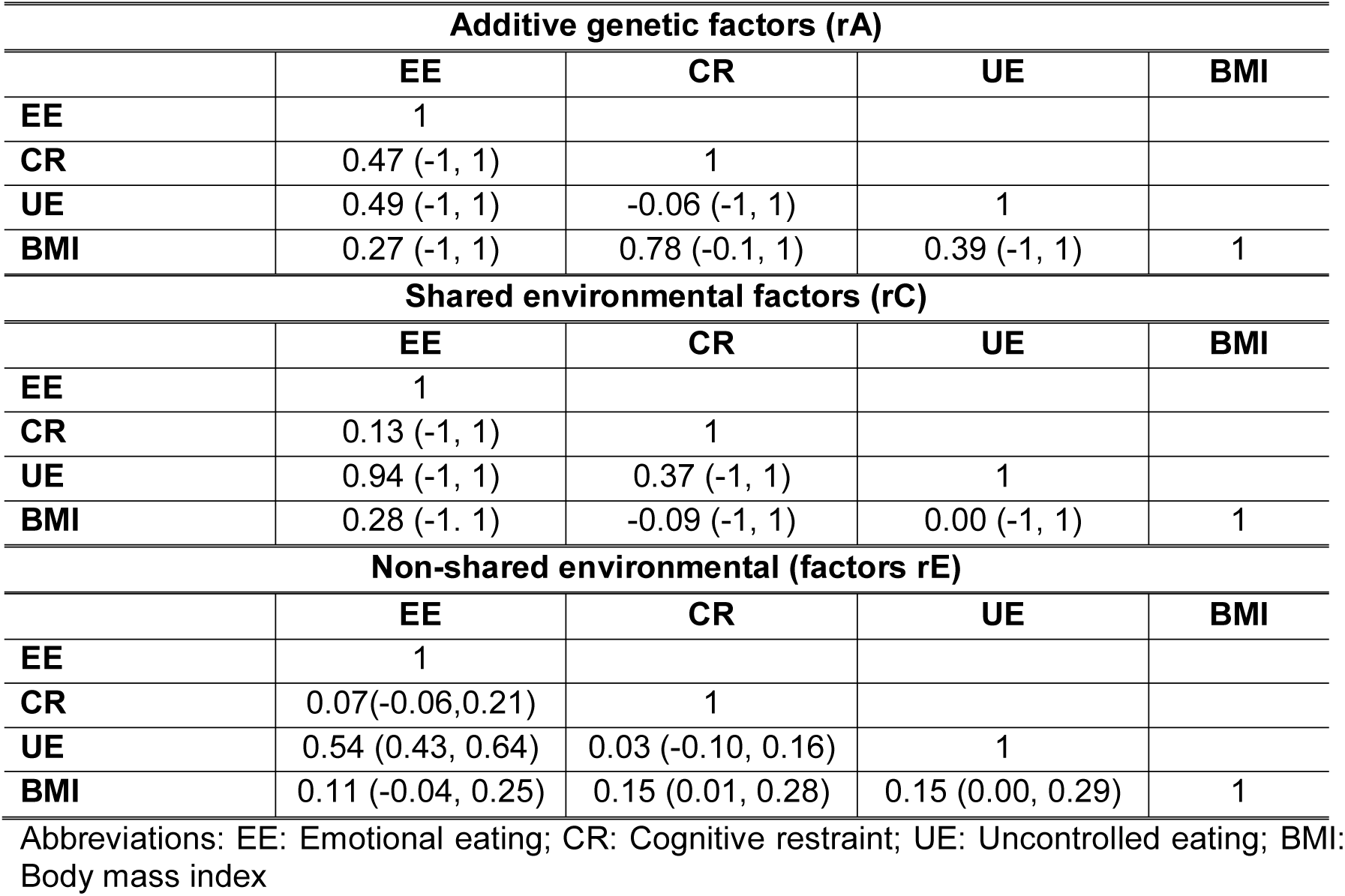
Results from multivariate ACE model, etiological correlations rA, rC and rE (95% Confidence intervals).

| **Additive genetic factors ($r_A$)** |  |  |  |  |
| --- | --- | --- | --- | --- |
|  | **EE** | **CR** | **UE** | **BMI** |
| **EE** | 1 |  |  |  |
| **CR** | 0.47 (-1, 1) | 1 |  |  |
| **UE** | 0.49 (-1, 1) | -0.06 (-1, 1) | 1 |  |
| **BMI** | 0.27 (-1, 1) | 0.78 (-0.1, 1) | 0.39 (-1, 1) | 1 |
| **Shared environmental factors ($r_C$)** |  |  |  |  |
|  | **EE** | **CR** | **UE** | **BMI** |
| **EE** | 1 |  |  |  |
| **CR** | 0.13 (-1, 1) | 1 |  |  |
| **UE** | 0.94 (-1, 1) | 0.37 (-1, 1) | 1 |  |
| **BMI** | 0.28 (-1, 1) | -0.09 (-1, 1) | 0.00 (-1, 1) | 1 |
| **Non-shared environmental (factors $r_E$)** |  |  |  |  |
|  | **EE** | **CR** | **UE** | **BMI** |
| **EE** | 1 |  |  |  |
| **CR** | 0.07(-0.06,0.21) | 1 |  |  |
| **UE** | 0.54 (0.43, 0.64) | 0.03 (-0.10, 0.16) | 1 |  |
| **BMI** | 0.11 (-0.04, 0.25) | 0.15 (0.01, 0.28) | 0.15 (0.00, 0.29) | 1 |

Abbreviations: EE: Emotional eating; CR: Cognitive restraint; UE: Uncontrolled eating; BMI: Body mass index

## Discussion

In summary, the present study explores the sources of variance and covariance of BMI and eating behaviors in a sample of adult women living in Murcia (Spain). Our results highlight the importance of environmental factors in the etiology of individual differences in eating behaviors and in their relationship with BMI. More specifically, our results suggest that the relationship between eating behaviors and BMI, at least in populations with these characteristics similar to ours, may be mainly driven by unique experiences rather than genetic factors, supporting the value of environmental factors as viable intervention targets in older cohorts.

Our findings are partially in line with previous twin studies of adult eating behaviors, confirming the importance of environmental factors in those behaviors (M. P. Herle et al., 2019; Keskitalo et al., 2008; Sung et al., 2010; Tholin et al., 2005). In addition, heritability of BMI was moderate and in line with previous work showing that genetic factors on BMI tend to increase during childhood (Haworth et al., 2008) and into adulthood (Min, Chiu, & Wang, 2013), but then decrease again in older adulthood (Nan et al., 2012). Notwithstanding, our study did not find convincing evidence of shared genetic etiology between BMI and any of the other eating behaviors, and confidence for all r_A_ estimates crossed zero. This is in contrast to findings from twin studies in infancy (C. H. Llewellyn et al., 2010b) and childhood (C. H. Llewellyn et al., 2014), which reported some shared genetic etiology between eating behavior and BMI. Moreover, in our sample, the relationship between eating behaviors and BMI was mainly explained by unique environmental factors, which is opposite to the findings by Keskitalo et al, the only other twin study that has explored this area. However, it should be remarked that their sample was quite different to ours, both in terms of geographical origin (i.e., UK and Finland) and age range (i.e., 17-82 years).

The following implications of this study might be considered: First, emotional eating and uncontrolled eating are highly positively correlated in our sample and share approximately half of the unique environmental factors of their underlying etiology. This might be interpreted as environmental triggers associated with emotional eating are also associated with uncontrolled eating. This is in line with previous research indicating that negative emotions (which are likely to be influenced by environmental factors) trigger overeating and binge eating behavior (Leehr et al., 2015). Second, individual differences in cognitive restraint are partially explained by shared environmental factors in our sample. Previous research has suggested that siblings and parents act as role models and inspire dietary behaviors, such as cognitive restraint (Coomber & King, 2008). This might imply that twins encourage each other to lose weight and constrain their food intake, resulting in higher estimates of shared environmental effects. This highlights the potential of including close family members or friends, when designing interventions aimed at weight management. Family wide interventions are common practice for children and adolescence (Berge & Everts, 2011) but might be equally important for older adults, especially when living in close-knit communities. For childhood and adolescent obesity, previous research has highlighted the important influence of siblings (Park & Cormier, 2018) and close friends (Salvy, Feda, Epstein, & Roemmich, 2017), but less is known about the influence of siblings on dietary behaviors in middle and late adulthood This rationale could extend to group interventions, which harness the dynamic interactions between group members to facilitate behavior change (Paul-Ebhohimhen & Avenell, 2009). Third, despite the established evidence of the strong genetic etiology of obesity (Locke et al., 2015; Min et al., 2013), our study highlights how, in this sample, environmental factors are important in explaining individual differences. This finding is key, as environmental interventions, such as calorie restriction, remain the most common strategies to lose weight. Eating behaviors, like emotional eating and uncontrolled eating, are positively correlated with BMI and are potential intervention targets to help people to reach and maintain healthy weight. Weight loss interventions, targeting eating behaviors specifically, are currently in development and have shown some promise (Hunot, Fildes, Croker, Johnson, & Beeken, 2016).

### Strengths and Limitations

As all other twin research, this study needs to conform to the assumptions underlying the twin method. ‘The Equal Environment Assumption’ (EEA) states that environmental exposures influencing the variation of a trait are unrelated to the zygosity of the twin pairs – i.e. that MZs and DZs share their environments to the same extent. A violation of the EEA could lead to an overestimation of the genetic contribution to variation. However, previous studies have confirmed the validity of the EEA in twin studies in general, as well as specifically in twin research studying eating behaviors (Klump, Holly, Iacono, McGue, & Willson, 2000). For this twin registry specifically, previous research has found no violation of EEA (Sánchez-Romera, 2013). Our sample included only women. Henceforth we were not able to investigate etiological differences between men and women. Previous twin studies have discussed these differences, e.g. a higher heritability of emotional eating in women (M. P. Herle et al., 2019). Furthermore, eating behaviors were self-reported, which might have introduced reporting bias. However, the Three Factor Eating Questionnaire is a widely used tool and has been validated against measures of food intake (de Lauzon et al., 2004) and correlates with BMI in the expected direction in this sample. An additional limitation is the reduced sample size, resulting in large confidence intervals, precluding a straightforward interpretation of the decomposition of the covariance.

The main strengths are the characteristics of the Murcia Twin Registry and the distinctiveness of the sample. Female participants in this study are in their perimenopausal years. They had on average high body weight and two thirds of the sample had either overweight or obesity. Instead of over representing wealthy and healthy individuals, this cohort is population-based and therefore representative of Spain / the Region of Murcia, with homogeneous and well-known geographic and cultural characteristics. This makes this cohort unique, as it gives insight into the relative influence of genetic and environmental factors in a sample considered to be at higher risk of developing overweight and obesity.

### Conclusions

Findings of this study suggest that eating behaviors are positively associated with BMI. Although limited in statistical power to identify other factors, non-shared environmental factors explain most of the individual differences and are also responsible for some of their association in our sample. Eating behaviors might be promising intervention targets, supporting individuals in reaching and maintaining healthy weight. Finally, twin studies are a powerful tool to study the influence of environmental factors when the trait of interest (BMI in our case) is clearly influenced by genetic factors.

## Data Availability

Data can be requested from the authors.
All analytical code is stored here:
https://github.com/MoritzHerle/The-role-of-the-environment-in-overweight-and-eating-behavior-variability

## Acknowledgments

The MTR is being funded by Seneca Foundation — Regional Agency for Science and Technology, Murcia, Spain (grants 03082/PHCS/05, 08633/PHCS/08, 15302/PHCS/10 and 19479/PI/14) and the Spanish Ministry of Science and Innovation (PSI2009–11560 and PSI2014-56680-R). Dr Herle is funded by fellowship from the Medical Research Council UK (MR/T027843/1). Dr Colodro-Conde is supported by a QIMR Berghofer fellowship.

## Author contributions

MH, LCC and JO designed the research. MH, JMV and JMO analyzed the data. MH wrote the first draft of the manuscript. All authors have read and approved the final article

## Declaration of interest

Authors report no conflict of interest.

## Notes

### Competing Interest Statement

The authors have declared no competing interest.

